# Wastewater surveillance of antibiotic resistant bacteria for public health action: Potential and Challenges

**DOI:** 10.1101/2024.03.31.24305136

**Authors:** Betsy Foxman, Elizabeth Salzman, Chelsie Gesierich, Sarah Gardner, Michelle Ammerman, Marisa Eisenberg, Krista Wigginton

**Affiliations:** Center for Molecular and Clinical Epidemiology of Infectious Diseases, Department of Epidemiology, School of Public Health, University of Michigan, Ann Arbor, Michigan, United States; Department of Civil and Environmental Engineering, College of Engineering, University of Michigan, Ann Arbor, Michigan, United States; Departments of Epidemiology, School of Public Health, Department of Mathematics and Center for the Study of Complex Systems, College of Literature, Sciences, and the Arts, University of Michigan, Ann Arbor, Michigan, United States

**Author notes:** All authors contributed equally to the work. Data-availability statement*: The data supporting the findings of this study are available within the article and/or its supplementary materials.

**Keywords:** surveillance, antibiotic resistance, wastewater epidemiology

## Abstract

Antibiotic resistance is an urgent public health threat. Actions to reduce this threat include requiring prescriptions for antibiotic use, antibiotic stewardship programs, educational programs targeting patients and healthcare providers, and limiting antibiotic use in agriculture, aquaculture, and animal husbandry. Wastewater surveillance might complement clinical surveillance by tracking time/space variation essential for detecting outbreaks and evaluating efficacy of evidence-based interventions; identifying high-risk populations for targeted monitoring; providing early warning of the emergence and spread of antibiotic resistant bacteria and identifying novel antibiotic resistant threats. Wastewater surveillance was an effective early warning system for SARS-CoV-2 spread and detection of the emergence of new viral strains. In this data-driven commentary we explore whether monitoring wastewater for antibiotic resistant genes and/or bacteria resistant to antibiotics might provide useful information for public health action. Using carbapenem resistance as an example, we highlight technical challenges associated with using wastewater to quantify temporal/spatial trends in antibiotic resistant bacteria (ARBs) and antibiotic resistant genes (ARGs) and compare with clinical information. While ARGs and ARBs are detectable in wastewater enabling early detection of novel ARGs, quantitation of ARBs and ARGs with current methods is too variable to reliably track space/time variation.

## Background

Wastewater surveillance for SARS-CoV-2 has proven benefit (1). Wastewater provides an estimate of the population prevalence of all SARS-CoV-2 cases - symptomatic and asymptomatic - and provides an early warning of the emergence and spread of new variants complementing data obtained from clinical surveillance. The value of wastewater surveillance has also been demonstrated for endemic diseases such as polio, influenza, norovirus and rotavirus (2–4). Biomarkers evaluating the amount of virus present in wastewater can be accurately quantified and normalized to account for the time-varying amount of human material in wastewater and the analytical recovery of the target from the wastewater. Wastewater surveillance provides timely, local information regarding the start of the respective seasons enabling implementation of vaccination (for influenza) and educational campaigns (for influenza, norovirus, and rotavirus) (5). For example, a recent study by Ammerman *et al*. used RT-ddPCR to evaluate wastewater surveillance of human norovirus (HuNoV GII); wastewater surveillance consistently coincided or lead syndromic surveillance for norovirus outbreaks. This finding suggests that wastewater surveillance is effective and able to inform public health interventions for human norovirus (6). Wastewater surveillance data for these viral pathogens are already available in local (e.g., https://um.wastewatermonitoring.dataepi.org) and national dashboards (e.g., https://data.wastewaterscan.org). The goal of this data-driven commentary is to assess whether wastewater surveillance for antibiotic resistant bacteria (ARBs) and antibiotic resistant genes (ARGs) will also provide useful information for public health action, complementing ongoing surveillance efforts.

Antibiotic resistance is an urgent public health threat. Each year in the United States more than 2.8 million antibiotic infections occur resulting in ∼35,000 deaths (7). Among the 35 countries in the Americas, an estimated 2 out of every 5 infection deaths were associated with antibiotic resistant bacteria (8). Actions to reduce this threat include requiring prescriptions for antibiotic use, antibiotic stewardship programs, educational programs targeting patients and healthcare providers, and limiting antibiotic use in agriculture, aquaculture, and animal husbandry. Lacking are population estimates of antibiotic resistance that are independent of the healthcare system: many ARBs and ARGs can be detected in the feces or on the skin of healthy humans. Wastewater surveillance for antibiotic resistance is feasible: ARBs and ARGs are excreted from feces, urine, nasal mucus, skin and sputum during all stages of infection and are detectable in wastewater (9). But to inform public health action, wastewater surveillance for ARBs and ARGs needs to provide information that complements ongoing clinical surveillance for antibiotic resistance, like the benefits that made wastewater surveillance for viruses valuable. This information would include: 1) establishing baseline levels which are essential for tracking time/space variation, detecting outbreaks and evaluating efficacy of evidence-based interventions; 2) identifying high-risk populations for targeted monitoring; 3) providing early warning of the emergence and spread of antibiotic resistant bacteria; and 4) identifying novel antibiotic resistant threats.

Extended spectrum beta lactamase (ESBL) producing Enterobacteriaceae and carbapenem resistant Enterobacteriaceae (CRE) are of particular concern because ESBL destroy beta-lactam antibiotics (e.g., penicillin and penicillin derivatives), and carbapenems are one of the few antibiotics effective against ESBL bacteria. The Enterobacteriaceae family includes three nosocomial pathogens, *Enterococcus faecium, Klebsiella pneumoniae*, and *Enterobacter spp.,* that are often resistant to multiple antibiotics, and several common foodborne pathogens, including *Escherichia coli*, Shigella, and Salmonella that are increasingly antibiotic resistant. In 2019, Enterobacteriaceae producing ESBL and CRE resulted in 197,400 cases and 9,100 deaths and 13,100 cases and 1,100 deaths, respectively in the United States (7).

Relative to humans, Enterobacteriaceae have very short generation times. The generation time of *E. coli*, for example, may be as short as 15 minutes. Spontaneous mutations from errors during DNA replication or exposure to mutagens occur at a rate between 1 in 10 million to 1 in 1 billion base substitutions per nucleotide per generation, but higher mutation frequencies have been reported for *E. coli* and Salmonella (10). The short generation time and large population size (39 trillion bacteria cells in each human (11)) virtually ensure that with the continued use of a given antibiotic, one or more human bacterial pathogens will become resistant to that drug. Requiring prescriptions for antibiotic use, antibiotic stewardship programs, and educational programs targeting patients and healthcare providers can lengthen the time until resistance appears. However, these prevention strategies reduce but do not eliminate the effects of antibiotic contamination of the environment.

Before penicillin was commercially available, urine from treated patients was collected and penicillin re-extracted for re-use, because penicillin (and other antibiotics) is poorly metabolized by animals (12). Therefore, it follows that when antibiotics are used clinically, in aquaculture, and animal husbandry, antibiotics absorbed into the bloodstream are excreted renally (via the urine) or via the liver (via the feces). Antibiotics that are not absorbed are excreted in the feces. Thus, as wastewater moves from homes and businesses via the sewers towards a wastewater treatment plant, the presence of antibiotics and chemicals can induce horizontal transfer of genes for antibiotic resistance (13,14).As ARGs and ARBs have been detected in wastewater worldwide (15), there has been an increased interest in the potential to use wastewater to detect and quantify ARGs and ARBs as a basis for public health action (16). Surveillance of wastewater prior to wastewater treatment has the potential to complement ongoing clinical surveillance for antibiotic resistance as wastewater detects ARBs and ARGs present among community members regardless of health status or use of healthcare.

Whether this potential is reachable is yet to be determined. A 2022 systematic review of 48 articles noted that most studies focused only on clinically relevant pathogens of fecal origin, which tend to have higher loads in wastewater facilitating detection, and that the fates of each bacterial species and ARG within wastewater varied. The authors also underscored that ARGs are found in animal hosts, symbiotic bacteria, and environmental sources (17). Therefore, community baselines will need to be species specific and factor in variability due to non-human sources. In a systematic review and qualitative synthesis of 33 studies that used a variety of detection methods, including culture, PCR and metagenomics, Chau *et al*. described concordance between estimates of human ARB prevalence in wastewater and clinical estimates (18). The authors identified high concordance of targeted ARGs (primarily CTX-M beta-lactamases) detected in selected bacteria (most often *E. coli*) found in wastewater and clinical samples. The four studies using metagenomics did not have extractable data, but only two appeared to find an association between ARBs detected in wastewater and humans. Both reviews noted that best practices for sampling and testing remain to be developed and the need for longitudinal studies.

Should these limitations be overcome, wastewater surveillance also may be useful for tracking antibiotic resistant foodborne illness. Many foodborne related pathogens have antibiotic resistance that contributes to the spread of antibiotic resistant genes (19). A significant subset of the three of the five commonly reported virulent foodborne pathogens—Salmonella (31%), *Campylobacter* (5%), and *Escherichia coli* O157:H7 (3%)—are antibiotic resistant (20). Currently, clinical surveillance in which health care providers submit reports to the Foodborne Disease Outbreak Surveillance System (FDOSS) (21) is the primary method for tracking foodborne illness. However, clinical surveillance often underestimates the true prevalence of foodborne illness and outbreaks often go undetected. Despite this underreporting, the Centers for Disease Control records 48 million illnesses, 128,000 hospitalizations, and 3,000 deaths that are reported each year in the United States (19). In principle, wastewater surveillance might lead to better estimates of the population burden of antibiotic resistant foodborne illnesses including the true case load, the temporality of outbreaks, and length of shedding (22). Further, improving surveillance for common foodborne-related pathogens would strengthen state and local health department capacity to investigate and report outbreaks, identify emerging food safety issues, and assess infection control interventions (21).

Given the ubiquity of foodborne bacteria in the environment and animals (including humans), it may be extremely difficult to implement wastewater surveillance to prospectively detect foodborne outbreaks (21,23). However, monitoring for foodborne bacteria with known genotypes (e.g., those with specific antibiotic resistances and virulence factors) may enable tracking of an ongoing foodborne outbreak and minimize false positives arising from commensal carriage, animal sources and foodwaste. Currently, whole genome sequencing is the most promising method for analyzing foodborne bacterial pathogens because it is the most specific and comprehensive method of identification and can link isolates from a similar source. Salmonella, *Campylobacter* colonize and *Escherichia coli* O157:H7 colonize food and other animals; foodborne outbreaks often follow consumption of uncooked eggs, milk, undercooked meat from food animals, food items contaminated by food or other animal feces. However, even with whole genome sequencing we cannot determine the pathogen host, making it impossible to assess if a given pathogen is human related prior to an outbreak (24,25).

Research is ongoing to use next generation sequencing to detect foodborne pathogens in wastewater. The challenges include low sensitivity and false positive results at low concentrations – problems which are likely to occur in wastewater samples (26). Additionally, time and resource consuming culturing methods must be in place to select for relevant pathogens before undergoing molecular and genomic characterization, which makes practical implementation for public health surveillance daunting (24). Real time detection technologies such as Nanopore Sequencing may be useful in overcoming this barrier, but further development and investigation is needed (27).

At the time of this writing, only two outbreaks of foodborne bacterial infections, both of salmonella, have been successfully surveilled in wastewater and reported in the literature. These two reports retrospectively detected two different salmonellosis outbreaks in Honolulu, Hawaii in wastewater samples from 2010-2011. Using genomic characterization, they were able to detect an increase in the serovar of salmonella related to an outbreak during that period. Additionally, they detected a resurgence of salmonella after the outbreak that clinical surveillance had missed (28,29). However, unlike viral pathogens, there are no reports of prospective wastewater surveillance being used to detect outbreaks of enteric bacterial pathogens.

In summary, ARBs and ARGs are detectable in wastewater but how to use that information for public health action is less clear. ARGs found in human pathogens also occur in bacteria found in animals and in the environment complicating source tracking. Variation in target material due to variability in flow rates and dilution make quantitation of colony counts from culture or gene copies from non-culture assessment difficult. There are few longitudinal assessments. Further, as most studies to date have targeted known human pathogens, there are few estimates of the occurrence of ARGs in other bacterial species. In the remainder of this commentary we use our studies of the detection of carbapenem resistant bacteria and genes to highlight technical challenges associated with using wastewater to quantify temporal/spatial trends in ARBS and ARGs and compare our findings with clinical information. We close with our assessment that -- at least with current technologies -- the primary value added of screening wastewater for ARBs and ARGs for direct public health action will be early detection of novel ARGs; quantitation with current methods is too variable to reliably track space/time variation

### Evaluating the Utility of Wastewater Surveillance for ARGs and ARBs

As a first step towards evaluating the utility of wastewater surveillance for ARGs and ARBs we used a laboratory pipeline enabling us to detect human pathogens with an antibiotic resistant phenotype to ARGs that code for that resistance, in this case, carbapenem resistance. This multistep laboratory pipeline includes filtering influent and suspending the filtrate in media containing the target antibiotic (Figure 1). The filtrate is next diluted and streaked on chromogenic agar, which gives a presumptive species identification. The species of each isolate is confirmed using MALDI-ToF (matrix assisted laser desorption ionization time of flight) and DNA is extracted and tested for the four targeted ARGs using qPCR– *bla*_KPC_, *bla*_NDM_, *bla*_OXA48_, and *bla*_VIM_. Because our pipeline directly measures phenotype, our results do not include ARGs from dead cells and also identifies intrinsically antibiotic resistant bacteria or resistance due to unknown or untargeted mechanisms. The presence of the four ARGs was determined in “wastewater microbiome” samples by extracting DNA from the filtered microbes prior to plating. These results represent a snapshot of ARGs on a given day from each plant but may include ARGs from dead cells.

**Figure 1:**
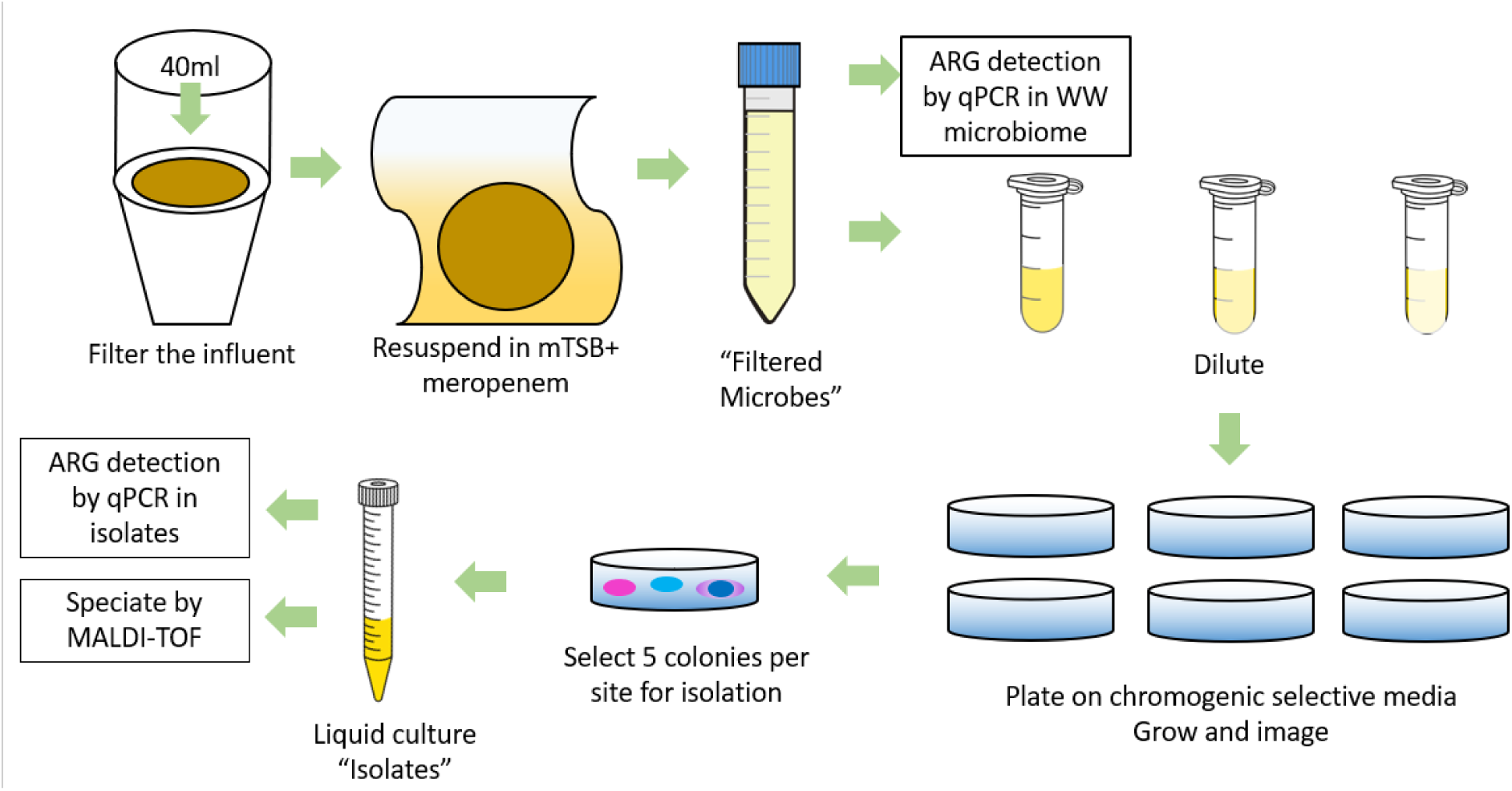
Laboratory pipeline for detecting and speciating antibiotic resistant human pathogens and identifying the genes coding for antibiotic resistance among the identified species. Filtered microbes captures the wastewater microbiome.

We used qPCR to directly detect carbapenemase genes from the filtered samples collected from the two wastewater treatment plants over the one-year period. *bla*_NDM_ positivity was verified by Sanger sequencing. We observed little or no variation in percent of carbapenemase genes observed over time of collection and results were similar between treatment plants. We detected *bla*_KPC_, *bla*_NDM_, and *bla*_OXA48_ in virtually all samples. *bla*_VIM_ was detected less frequently, with similar patterns of detection in the two wastewater treatment plants (See Figure S1 in Appendix).

From the filtrate collected from two wastewater treatment plants between June 2022 and June 2023 and plated on chromogenic media selective for carbapenem, we picked 672 bacterial colonies for further testing. Approximately 20% (134 of 672) of colonies picked for isolation did not grow further, highlighting the need for more extensive methods to elicit growth and suggesting that when using media that is enriched for Enterobacteriaceae there may be proximity-dependent and temporary sharing of ARGs. Of the 467 isolates with qPCR results for all 4 carbapenemase genes 395 could be speciated by MALDI (Figure 2).

**Figure 2:**
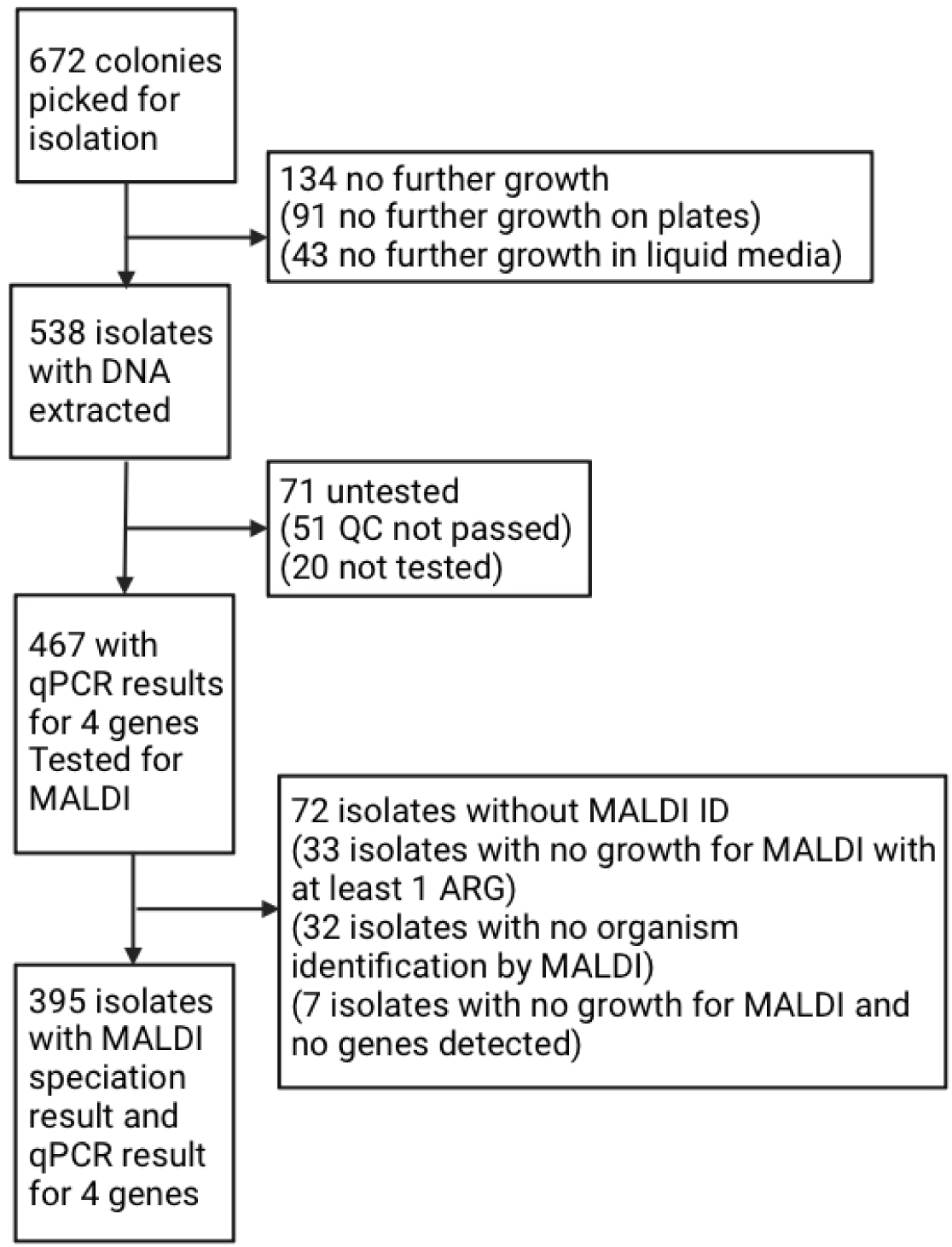
CONSORT diagram detailing wastewater isolates detection and processing. (MALDI= matrix assisted laser desorption ionization time of flight; QC= quality control).

Among the 395 isolates we identified 53 species: 11 were known pathogens and multiple species fit within the *Enterobacter cloacae* complex or the *Citrobacter freundii* complex. We most frequently detected human pathobionts *Klebsiella pneumoniae*, *Enterobacter cloacae* complex, *Klebsiella oxytoca* and *Citrobacter freundii* complex. One or more known carbapenemase genes were detected in virtually all isolates: 90% had *bla*_KPC_, 15% *bla*_NDM_, and 12% *bla*_OXA-48_ (data note shown). We did not detect *bla*_VIM_ in any of the isolated species. Using our pipeline, we were able to detect the presence of four known clinically relevant carbapenem resistant genes in identified bacterial pathogens within 4 to 5 days following sample receipt.

We compared our results from wastewater surveillance of samples from southeastern Michigan with clinical surveillance data for CRE (n=275) collected by the Michigan Department of Human Health and Services (30) for the entire State of Michigan over the same time period (Figure 3; see Figures S2 and S3 for other clinically relevant species, and Table S1 in appendix for listing of all species identified). There were some similarities and notable differences between the frequency of carbapenem resistance mechanisms identified in the most detected clinical isolates from the entire State and the same species detected in our two wastewater treatment plants (Figure 3). Although the frequency of KPC as a single resistance mechanism in *Klebsiella spp*. was similar between clinical and wastewater isolates (78% vs. 82%, respectively), in clinical isolates NDM and OXA-48 most often occurred alone; in wastewater NDM was only found in the presence of KPC or OXA-48 or both. The pattern was similar for *E. coli* and *Enterobacter spp.* although for *Enterobacter spp.* in a small percentage (3%) of wastewater isolates NDM was the sole resistance mechanism. Further, for *E. coli* and *Citrobacter freundii* OXA-48 was only detected together with KPC in wastewater (Figure 3). Higher levels of phenotypic-only resistance were seen in the clinically relevant species that were only detected in wastewater: Aeromonas species, *Pseudomonas aeroginosa*, (Figure S3) and the “Other species” category (Figure S2), suggesting other mechanisms of resistance being more prevalent in wastewater than in clinical samples.

**Figure 3:**
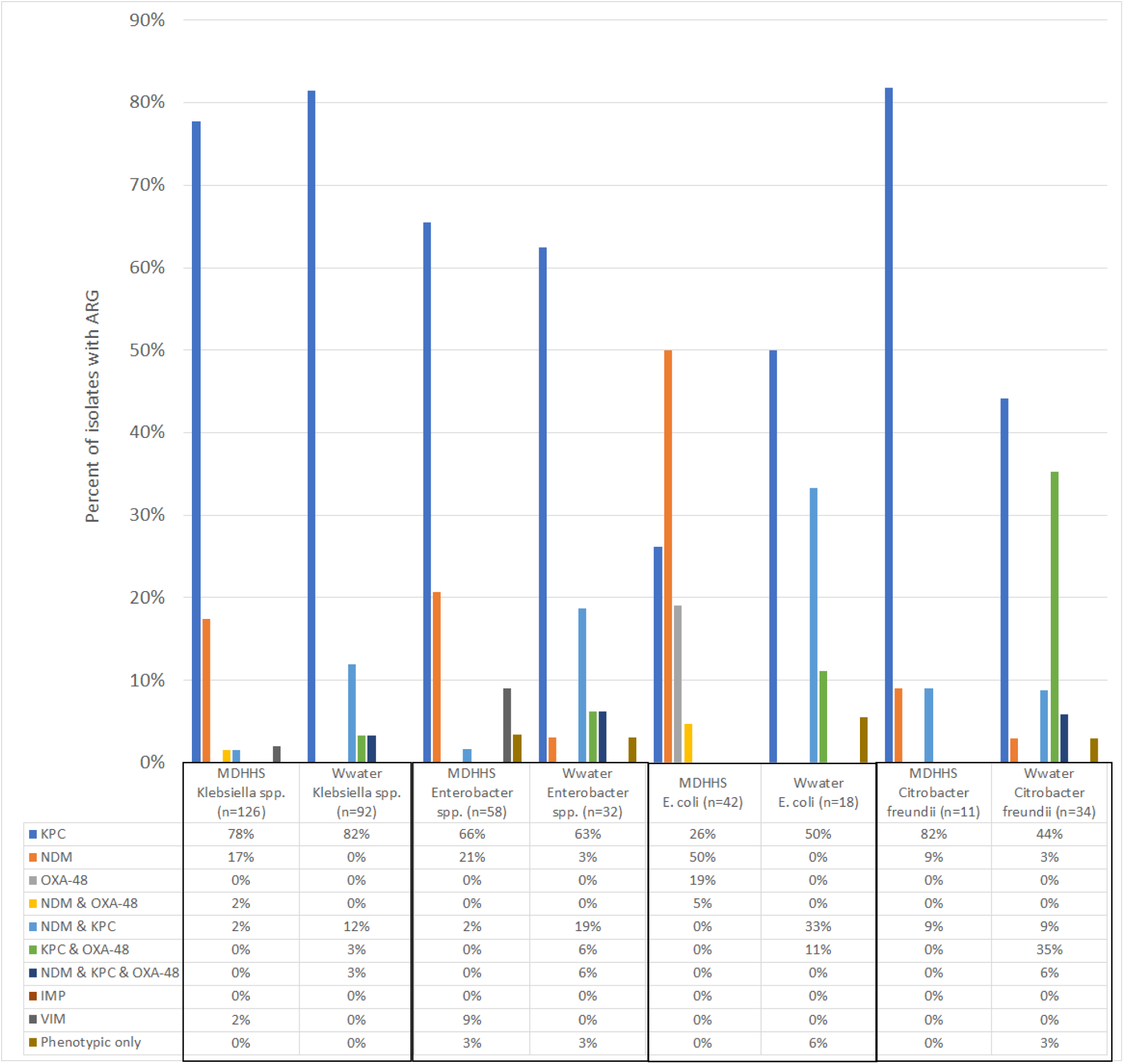
Percent of carbapenem-resistant *Klebsiella spp., Enterobacter spp*., *Escherichia coli* and *Citrobacter freundii* clinical isolates and wastewater-detected isolates by carbapenem genes (*bla*_KPC_, *bla*_NDM_, *bla*_OXA48_ and *bla*_VIM_). Clinical cases (n=275; 268 were typed for CRE genes) reported to the Michigan Department of Health and Human Services (MDHHS) in 2022 (30); wastewater (Wwater) isolates samples from two wastewater treatment plants between June 6, 2022 and June 12, 2023. Wastewater samples were collected weekly. See Supplement for all carbapenem-resistant species.

## Discussion

Bacteria are different than virus. Unlike the viral pathogens targeted for wastewater surveillance whose primary reservoir is humans, human bacterial pathogens like the ESKAPE pathogens (*Enterococcus faecium, Staphylococcus aureus, Klebsiella pneumoniae, Acinetobacter baumannii, Pseudomonas aeruginosa,* and *Enterobacter species*) – nosocomial pathogens that exhibit multidrug resistance and virulence – are found in non-humans and/or can live in the environment (Figure 4). Further, most human bacterial pathogens are pathobionts, that is, they can colonize humans (commensals or asymptomatic carriage) for extended periods only causing symptoms (disease) under certain circumstances. Methicillin resistant *Staphylococcus aureus* (MRSA) is just one example of many.

**Figure 4:**
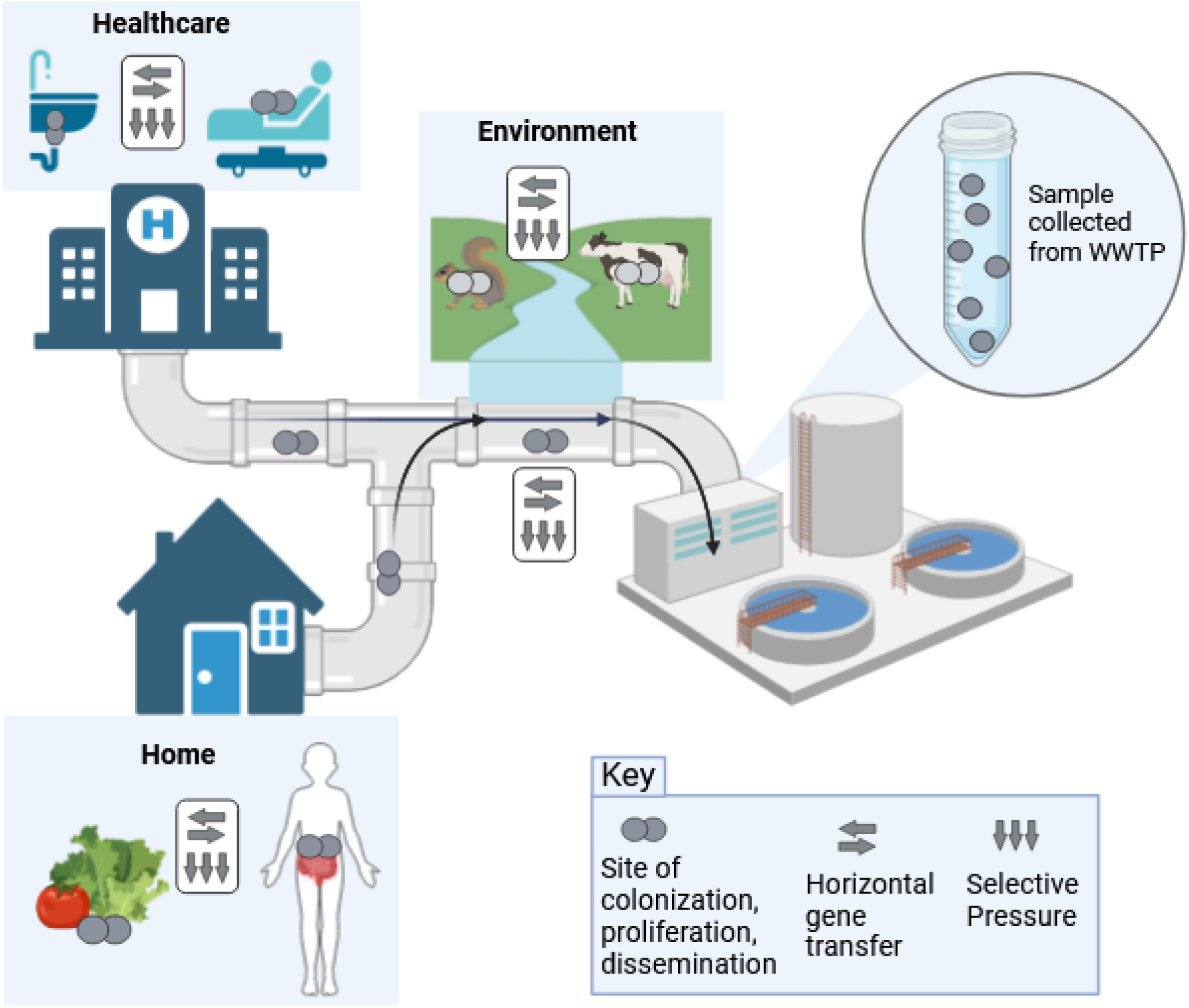
Sources of antibiotic resistant bacteria (ARBs) and antibiotic resistant genes (ARGs) in wastewater. ARBs found in humans, food, animals, agriculture, pipes, and sinks shed into wastewater. Within the wastewater ARGs can transfer between bacteria (horizontal gene transfer).

Our results demonstrate that ARGs can be detected in wastewater and linked to specific ARBs relatively quickly and that similar ARGs are detected in ARBs as in clinical surveillance. However, an additional comparison of our findings with literature and clinical surveillance is illuminating. A 2023 report estimated that in 2019 among 35 countries in the Americas there were 114,000 deaths associated with carbapenem resistant infections. Most of these deaths were attributed to infections with the following carbapeneum-resistant bacterial species (number of deaths): *Acinetobacter baumannii* (36,800), *Pseudomonas aeruginosa* (33,200), *Streptococcus pneumoniae* (18,100) and *K. pneumoniae* (16,500) (8). With our pipeline we identified only one *A. baumannii* and did not detect *S. pneumoniae* – but we did not use growth conditions specific for *S. pneumoniae*. We found one study that detected *Streptococcus spp*. and *Acinetobacter spp*. in wastewater; this study used 16S rRNA sequencing and did not resolve to the species level (31). We also found only one study that isolated *A. baumannii* from wastewater. Their pipeline started with 1 liter of influent which was serially diluted 10-fold and used CHROMagar to detect *A. baumannii,* then *c*arbapenemase genes were detected using qPCR (32).

Monitoring wastewater influent for new ARGs might alert clinicians that a current antibiotic may soon become less effective. Of the four carbapenem resistant genes we monitored over a seven-month period, only *bla*_VIM_ –which is also the newest– showed any variation in prevalence over time in the wastewater microbiome samples. While this suggests that only emerging ARGs show significant variation in prevalence over time and space, this remains to be determined.

Normalization is necessary to account for time-varying amounts of human material in wastewater and the analytical recovery of the target from the wastewater. The amount of material can vary with changes in flow volume, dilution from precipitation and variation in human waste input (toilet flushes). Although for our protocol we filtered a standard volume of water, the biomass within varied from sample to sample. Several different human biomarkers have been used to normalize estimates of SARS-CoV-2 from wastewater, e.g., pepper mild mottle virus, crassphage, and caffeine (33). Unfortunately, although human biomarkers are useful in the case of SARS-CoV-2 and other viruses, their utility for ARBs and ARGs is less certain because ARBs and ARGs have multiple non-human sources making it difficult to determine whether the ARBs and ARGs detected come from human or environmental sources. This poses a major challenge to using wastewater to estimate population prevalence or for monitoring trends over time – even if analyses account for changes in flowrates due to precipitation: increases of precipitation may increase the ARBs and ARG from environmental sources (e.g., from animal fecal matter and soil) in an unpredictable fashion.

Focusing wastewater surveillance on ARGs instead of ARBs also is problematic. ARGs are detectable in food (34,35), food animals, and in the biofilms found in sinks and pipes (36) [Figure 1]. ARGs can be transmitted between bacteria of the same or different species by bacterial pathogens (phage) or via horizontal gene transfer. Bacteria can also acquire ARBs by transformation, the uptake of environmental ARGs. *In vitro* experiments suggest that susceptible bacteria in the wastewater stream may acquire ARGs prior to wastewater treatment (13). Also, the same ARG sequence can be detected in multiple bacterial species isolated from different animals and environments (37). Thus, detection of ARGs in wastewater surveillance detects ARG presence, but gives little information about ARG occurrence in human pathogens or prevalence in human populations.

Further, the proportion of bacteria in influent that is from genera containing human pathogens is low. A 2019 study by Limayem *et al*. (31) used 16S rRNA sequencing to identify the bacterial genera found in a 1.5 liter sample from a single wastewater treatment plant. In this sample, 6.4% of the 195,967 reads were of the genus *Enterococcus* 0.063%, 0.004% *Staphylococcus*, 0.038% *Klebsiella*, *0.968% Acinetobacter, 21.356% Pseudomonas, 6.445% Streptococcus*, and 0.028% *Escherichia* (31). Note that there are over 50 species in the genus *Streptococcus* of which 10 are considered clinically relevant. The genera *Staphylococcus, Enterococcus* and *Pseudomonas* also include many species but only a small proportion are clinically relevant. The low frequencies of taxa of interest increase potential for error and missed or false detection.

Nonetheless, ARG detection is not without value for public health purposes, as it can be used to identify whether an ARG has been introduced locally. Many ARGs emerge from countries with high rates of unregulated antibiotic use in humans and animals and then spread globally. Therefore, identifying ARGs in an agricultural area without clinical cases might be informative for source tracking. However, beyond source tracking it is unclear that detection in wastewater would be value added over data provided from clinical surveillance for public health decision making. If a new ARG is detected in wastewater prior to clinical identification, it seems unlikely to lead to any additional interventions beyond notification that a new ARGs has been discovered until identified locally among patients.

Although not the focus of this commentary, there may be value in surveillance of outflow from wastewater treatment plants to inform OneHealth assessments of the risks of antibiotic resistance on environmental ecosystems (38). Within wastewater treatment plants there is a mixing of bacteria, ARBs, ARGs and antibiotics, providing a potential hotspot for horizontal gene transfer (39) and selection for antibiotic resistance that is discharged into the environment.

It also has been suggested that it might be beneficial to monitor the effluent from hospitals, longterm care facilities or other venues where there is high frequency of antibiotic use. Based on our experience with sampling of these types of facilities for SARS-CoV-2, we perceive some potential challenges. First, access to the effluent may be challenging due to placement of sewer lines and manhole covers limiting testing to grab samples. Second, ARBs and ARGs colonize hospital sink traps and these are released into the hospital effluent. Therefore levels in wastewater may not accurately reflect human colonization or disease (40); this is likely true for other healthcare facilities. Third, the smaller population sizes make estimates unstable and limitations in quantitation make it difficult to determine if detection represents single or multiple individuals. Ability for Public Health officials to act on information acquired without benefit of consent may be fraught.

## Summary and Conclusions

It is our assessment that the intrinsic features of human bacterial pathobionts and how bacteria acquire and exchange ARGs generally limits the utility of using wastewater surveillance for ARBs and ARGs to inform public health action such as triggering an outbreak investigation, or to complement ongoing clinical surveillance by identifying areas at high risk of antibiotic resistant infection. Several factors complicate quantitative estimation: bacteria proliferate and exchange ARGs outside of living hosts, qPCR and sequencing does not distinguish between viable and non-viable organisms, and multiple steps are required for laboratory processing, *e.g*. filtration, growing on selective media. Although it is feasible to detect the presence of emerging ARGs in wastewater, whether the information will lead to earlier or additional public health action beyond that suggested by existing monitoring systems is uncertain. Technological innovations, such as long read sequencing of specific bacteria species directly from wastewater (without culture) and focusing on obligate human pathogens, e.g., such as *Haemophilus influenzae*, *Helicobacter pylori*, *Neisseria gonorrhoeae*, *Neisseria meningitidis*, *Mycobacterium leprae*, *Salmonella typhi*, *Streptococcus pneumoniae*, *Streptococcus pyogenes*, *Vibrio cholerae* and *Treponema pallidum* (41), would alleviate the problem of determining if a target ARG came from a specific bacterial species, but would not alleviate other problems of interpretation: specifically, whether the bacterium came from a human and whether the ARG was acquired post shedding from the human once it entered the wastewater stream.

In summary, it is possible to detect ARGs and ARBs in wastewater, but difficult to use the results to estimate population prevalence and to monitor trends over time and space. The difficulties arise because human pathogens occur at relatively low prevalence in wastewater, and many are shed from non-human sources.

## Data Availability

All data produced in the present work are contained in the manuscript.

## Acknowledgements

Preprint information: posted on medRxiv (27June2024). The authors claim no conflicts.

## SUPPLEMENTAL MATERIAL

**Figure S1:**
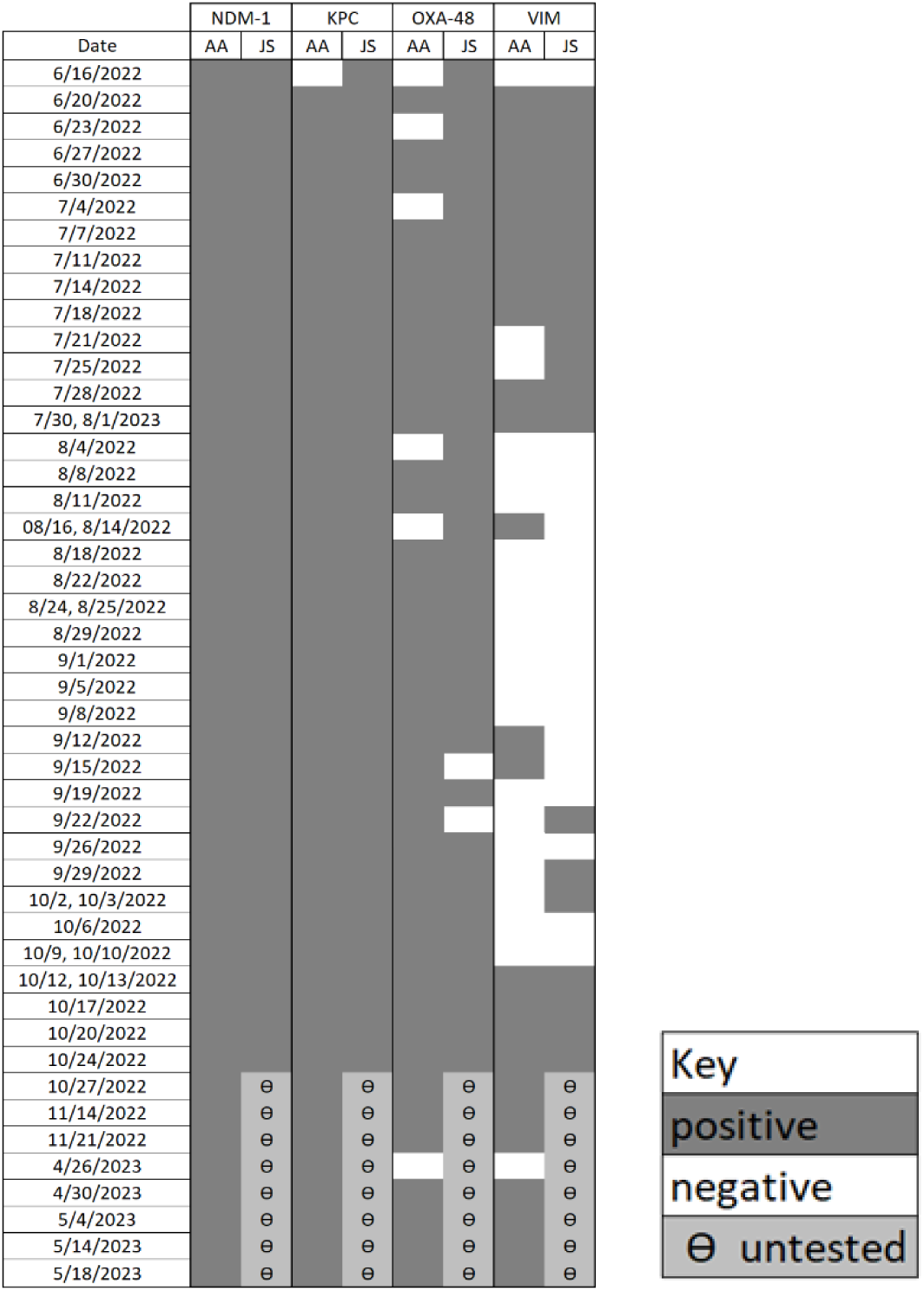
Presence of carbapenemase genes *bla*_VIM_, *bla*_KPC_, *bla*_NDM_, and *bla*_OXA48_ detected from filtered wastewater microbiome samples collected weekly from two Southeastern Michigan wastewater treatment plants (AA=Ann Arbor, JS=Jackson) between June 2022 and June 2023. Filtered microbes from the same day, or approximate same date, were tested for each ARG. (Stopped detection of samples from Jackson treatment plant in October 2022, because no longer informative).

**Table S1:**
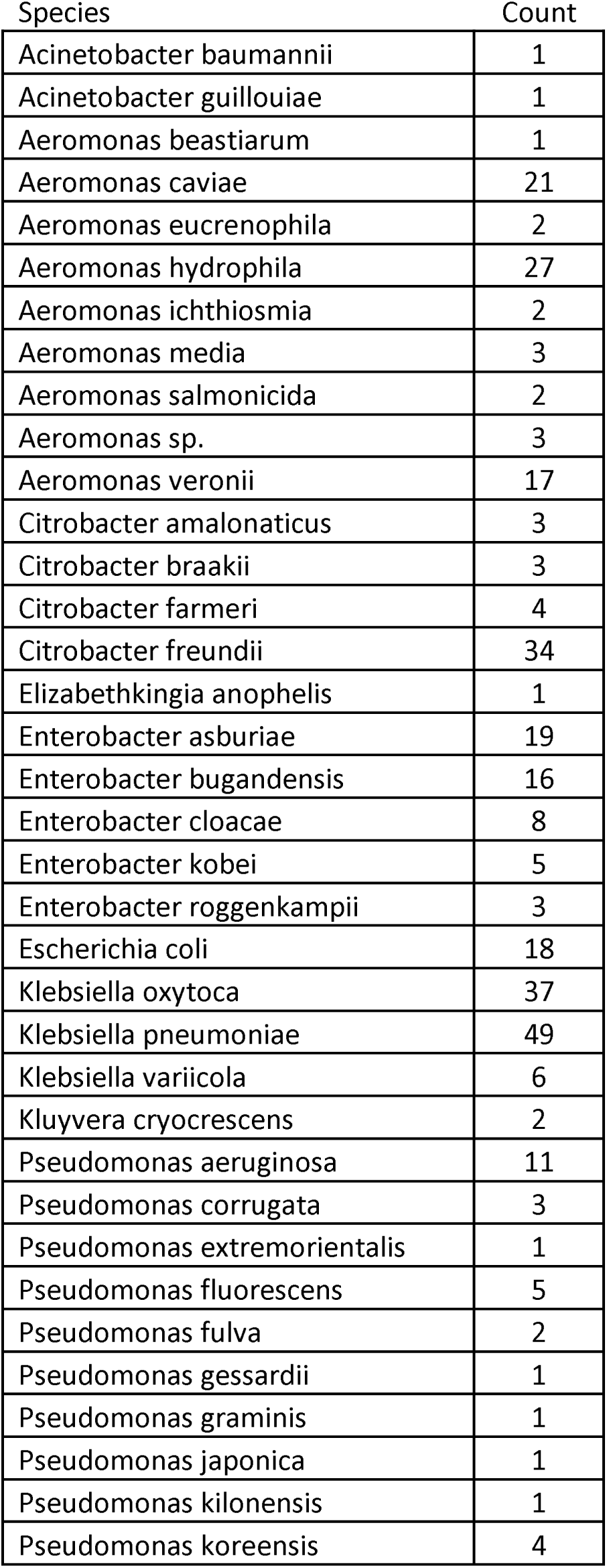
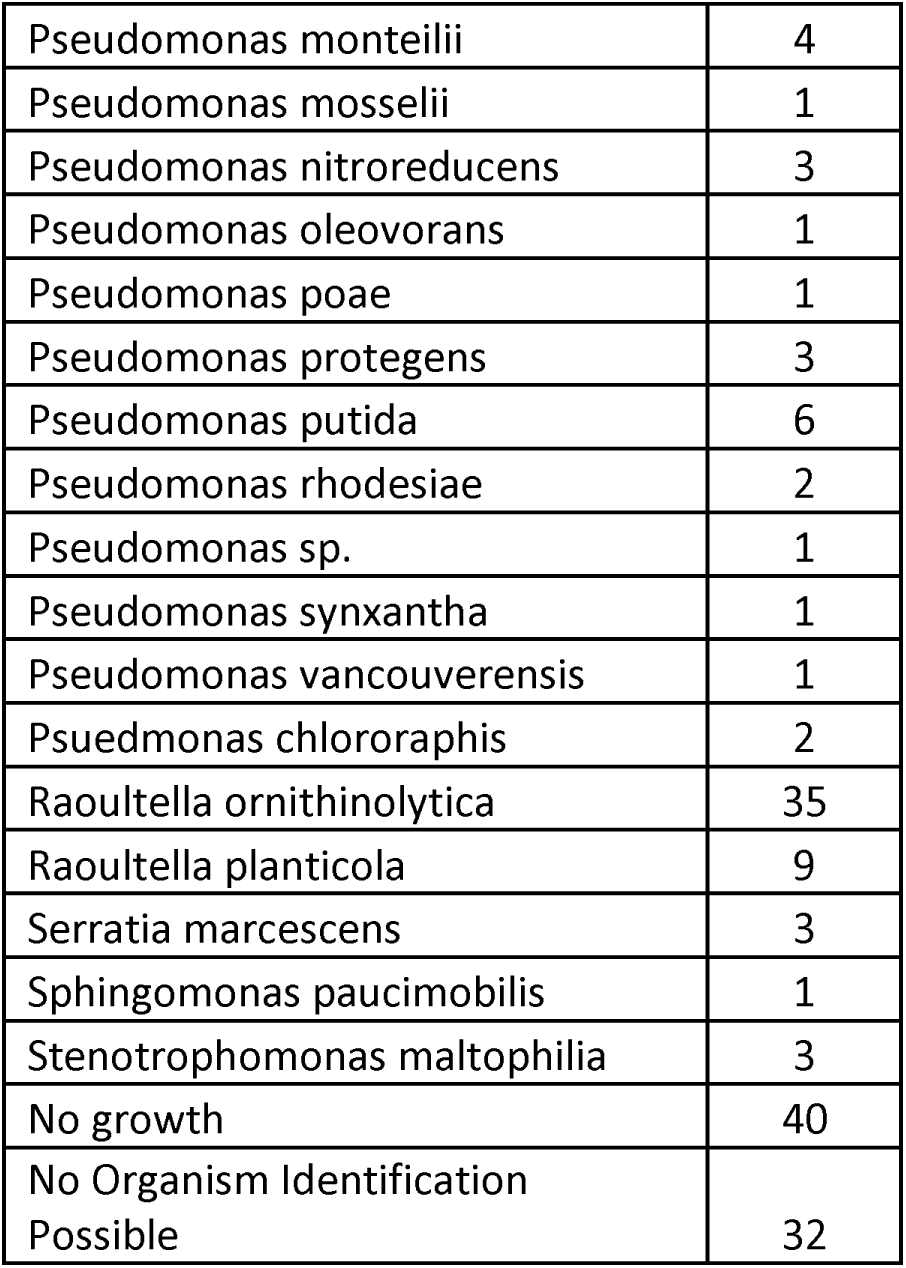
Frequency of all identified isolates by species detected from filtered wastewater microbiome samples collected weekly from two Southeastern Michigan wastewater treatment plants between June 2022 and June 2023.

**Figure S2:**
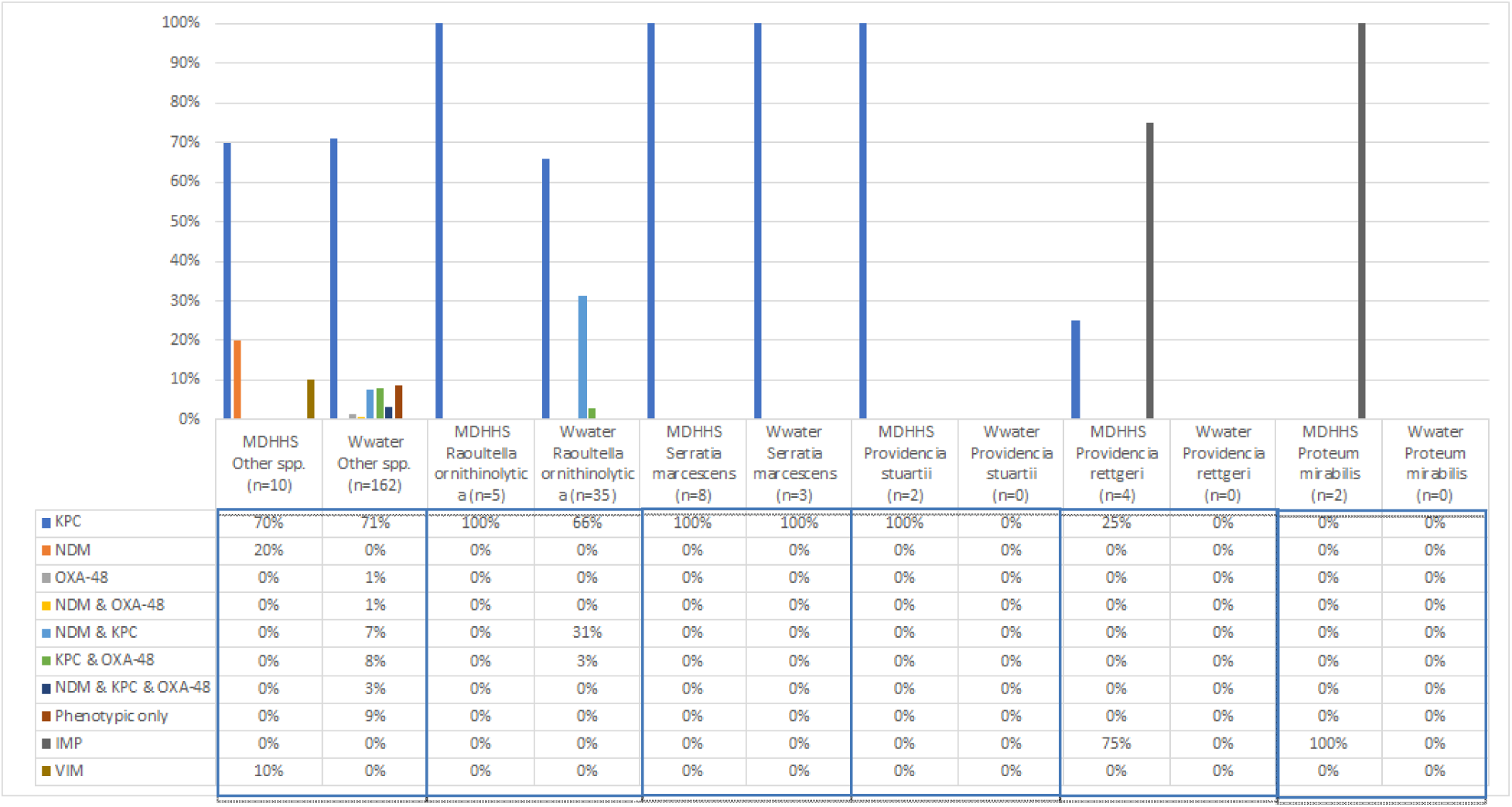
Percent of carbapenem-resistant “Other spp.” (species), *Raoultella ornithinolytica*, *Serratia marcescens*, *Providencia stuartii*, *Providencia rettgeri*, and *Proteum mirabilis* clinical isolates and wastewater-detected isolates by carbapenem genes (*bla*_KPC_, *bla*_NDM_, *bla*_OXA48_ and *bla*_VIM_). Clinical cases reported to the Michigan Department of Health and Human Services (MDHHS) in 2022; wastewater (Wwater) isolates samples from two wastewater treatment plants between June 6, 2022 and June 12, 2023. Wastewater samples were collected weekly. For Wwater isolates, “Other spp.” includes isolates that were tested for qPCR and unable to be identified by MALDI.

**Figure S3:**
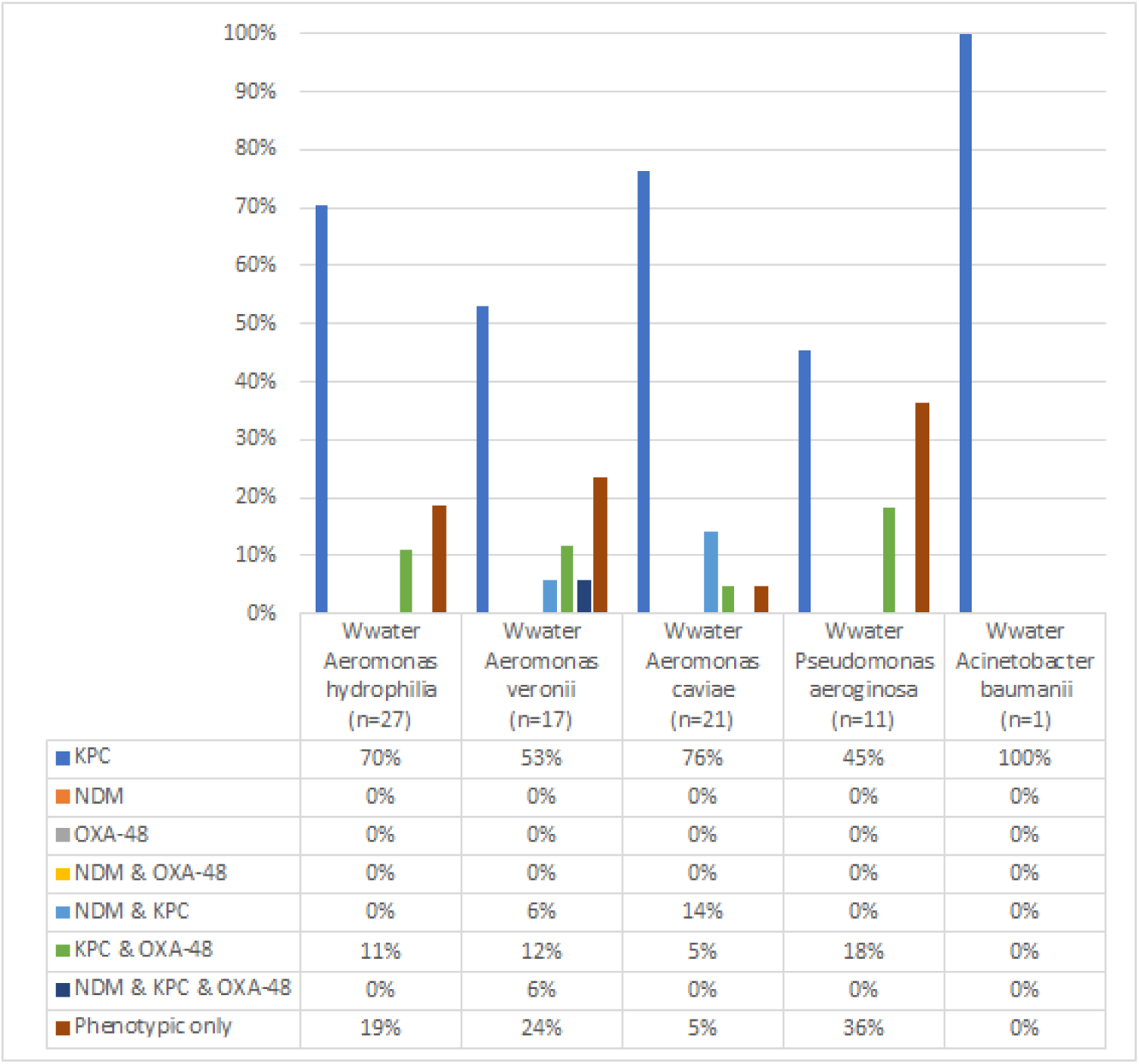
Percent of carbapenem-resistant *Aeromonas hydrophilia*, *Aeromonas veronii*, *Aeromonas caviae*, *Pseudomonas aeroginosa*, and *Acinetobacter baumanii* wastewater-detected isolates by carbapenem genes (*bla*_KPC_, *bla*_NDM_, *bla*_OXA48_ and *bla*_VIM_). Wastewater (Wwater) isolates samples from two wastewater treatment plants between June 6, 2022 and June 12, 2023. Wastewater samples were collected weekly.

## Notes

*Funding information:* This work was supported by the Michigan Department of Health & Human Services through the Michigan Sequencing Academic Partnership for Public Health Innovation and Response (MI-SAPPHIRE) grant (S MA-2022 (ELCEDE-UM) 10/01/2021 – 07/31/2024, (BF mPI), and the wastewater surveillance program (SEWER network grant, KRW and MCE co-PIs). The funders had no role in study design, data collection and analysis, decision to publish, or preparation of the manuscript.

### Competing Interest Statement

The authors have declared no competing interest.

### Funding Statement

This work was supported by the Michigan Department of Health & Human Services through the Michigan Sequencing Academic Partnership for Public Health Innovation and Response (MI-SAPPHIRE) grant (S MA-2022 (ELCEDE-UM) 10/01/2021 to 07/31/2024, (BF mPI), and the wastewater surveillance program (SEWER network grant, KRW and MCE coPIs). The funders had no role in study design, data collection and analysis, decision to publish, or preparation of the manuscript.

### Author Declarations

The used only openly available human data that were originally located at https://www.michigan.gov/mdhhs/-/media/Project/Websites/mdhhs/HAI-SHARP/CRE/CP-CRE-Surveillance-Report-2018-2022-Final.pdf?rev=b5ee21278e6c4c1c91e7ce116bf25403&hash=5AE501DE9961C2B742D69CD5CA453B16 (Accessed February 28, 2024)

### Summary of Updates

Revision reorganizing the presentation for clarity and elaborates the discussion.

